# Sales of over-the-counter products containing codeine in 31 countries, 2013-2019: a retrospective observational study

**DOI:** 10.1101/2021.04.21.21255888

**Authors:** Georgia C Richards, Jeffrey K Aronson, Brian MacKenna, Ben Goldacre, FD Richard Hobbs, Carl Heneghan

## Abstract

**Introduction:** Opioid prescribing trends have been investigated in many countries. However, the patterns of over-the-counter purchases of opioids without a prescription, such as codeine combinations, are mostly unknown.

**Objective:** We aimed to assess national sales and expenditure trends of over-the-counter codeine-containing products purchased in countries with available data over six years.

**Methods:** We conducted a retrospective observational study using electronic point-of-sale data from the human data science company, *IQVIA*, for countries that had such data, including Argentina, Belgium, Brazil, Bulgaria, Canada, Croatia, Estonia, Finland, France, Germany, Greece, Ireland, Italy, Japan, Latvia, Lithuania, Mexico, The Netherlands, Poland, Portugal, Romania, Russia, Serbia, Slovakia, Slovenia, South Africa, Spain, Switzerland, Thailand, the UK, and the USA. We calculated annual mean sales (dosage units per 1000 of the population) and public expenditure (GBP, £ per 1000 population) for each country between April 2013 and March 2019 and adjusted for data coverage reported by *IQVIA*. We quantified changes over time and the types of products sold.

**Results:** 31.5 billion dosage units (adjusted: 42.8 billion dosage units) of codeine, costing £2.55 billion (adjusted: £3.68 billion), were sold over-the-counter in 31 countries between April 2013 and March 2019. Total adjusted sales increased by 11% (3911 dosage units/1000 population in 2013 to 4358 in 2019) and adjusted public expenditure increased by 72% (£263/1000 in 2013 to £451/1000 in 2019). Sales were not equally distributed; South Africa sold the most (36 mean dosage units/person), followed by Ireland (30 mean dosage units/person), France (20 mean dosage units/person), the UK (17.2 mean dosage units/person), and Latvia (16.8 mean dosage units/person). Types of products (n=569) and formulations (n=12) sold varied.

**Conclusion:** In many parts of the world, substantial numbers of people may be purchasing and consuming codeine from over-the-counter products. Clinicians should ask patients about their use of over-the-counter products, and public health measures are required to improve the collection of sales data and the safety of such products.

**Study protocol pre-registration:** https://osf.io/ay4mc

The pre-print version of this work is available on medRxiv:

https://doi.org/10.1101/2021.04.21.21255888

**Key points:**

- Codeine is one of the most accessible pain medicines available worldwide, yet data on its use as an over-the-counter drug has been limited.
- We found that total sales and expenditure of over-the-counter products containing codeine increased from April 2013 to March 2019, but there was substantial variation in mean sales between countries and the coverage of data reported by *IQVIA*, with South Africa, France, Japan, the UK, and Poland accounting for 90% of all sales data.
- In countries with access to over-the-counter codeine products, sales data should be collected, made available, and reviewed to inform regulatory decisions and public health measures to ensure safety.

## 1 Introduction

Prescribing patterns of opioids are documented in many countries [1–4]. However, opioids such as analgesic combinations containing codeine can be purchased over-the-counter (OTC) without a prescription or consultation with a doctor or prescriber in most countries. As the access to data on sales of OTC medicines has been limited, previous research on the use of non-prescribed codeine has relied on case reports [5,6], self-reported questionnaires [7–10], qualitative studies [11–13], and data from poisons centres, hospital admissions, or coronial systems [14–16]. Since codeine is one of the most accessible opioids worldwide, an analysis of its use is needed to gauge a more robust understanding of opioid use globally.

Codeine (3-methylmorphine) is used for its analgesic, antidiarrheal, and antitussive effects [17–19]. It is often combined with other analgesics, such as paracetamol, and non-steroidal anti-inflammatory drugs (NSAIDs), such as ibuprofen. These combinations have greater efficacy than codeine alone [17,20,21]. But most clinical trials testing the efficacy of codeine have used high doses (25–90 mg), which are not available OTC [20–22]. A Cochrane overview of systematic reviews on oral OTC analgesics for acute pain found no studies or data that could be extracted on combinations of analgesics containing low doses of codeine [22]. A systematic review of the efficacy and safety of low-dose (≤30 mg) codeine included ten RCTs [23]. It reported low-to moderate-quality evidence that combination products of low-dose codeine provided little to moderate pain relief for acute and chronic pain conditions in the short term [23]. In observational studies, products containing codeine have been associated with dependence, misuse, death, and collateral toxicity from combinations with paracetamol and ibuprofen [15,24].

Regulation of codeine-containing products varies worldwide, making it difficult to estimate how much they are used [25]. Under the 1961 Single Convention on Narcotic Drugs, codeine is a Schedule III drug [26]. Drugs in this Schedule reportedly “are not liable to abuse and cannot produce ill effects”, and thus it is not mandatory to report data on their consumption to the International Narcotics Control Board (INCB). In a report presented at the WHO’s Expert Committee on Drug Dependence in October 2019, reviewing codeine formulations listed in Schedule III, the INCB reported a 64% increase in demand for codeine in the last decade [25]. Governments can also regulate codeine; for example, France (July 2017) and Australia (February 2018) have reclassified codeine to prescription-only [27,28]. A review of OTC codeine regulations in the European Union showed that more than half of member countries did not permit OTC sales of codeine as of March–August 2014 [29]. Studies have analysed the consumption of OTC cough syrup containing codeine in Taiwan [30] and the impact of rescheduling codeine to prescription-only in Australia [31,32], but the sales of OTC codeine products in other countries remains unknown. We aimed to assess trends in the sales and expenditure of products containing codeine sold OTC in countries with available data.

## 2 Methods

### 2.1 Design and data source

We conducted a retrospective observational study using consumer health sales data from *IQVIA* in the UK [33], which has previously been used in observational research on a range of medications [34–36]. The data included products sold OTC that contained codeine for adults, classified by IQVIA as pain relief or cough products. The data were collected using scan track barcodes from electronic point-of-sale (EPoS) store data in 31 countries: Argentina, Belgium, Brazil, Bulgaria, Canada, Croatia, Estonia, Finland, France, Germany, Greece, Ireland, Italy, Japan, Latvia, Lithuania, Mexico, The Netherlands, Poland, Portugal, Romania, Russia, Serbia, Slovakia, Slovenia, South Africa, Spain, Switzerland, Thailand, the UK, and the USA. The authors did not select these countries; they requested global sales data from *IQVIA* and received data for “all countries for which data is available” according to *IQVIA* at the time of data extraction (16 September 2019).

We received information about *IQVIA*’s coverage of data (Table S1 in Supplement 1) and quarterly sales from 1 April 2013 to 31 March 2019 in three types of measurements (1) numbers of packs and bottles (liquids) sold; (2) number of tablets or millilitres (mL) of liquids sold; and (3) dosage units. For our analysis, we used dosage units as this allows liquid and solid forms to be combined. Dosage units were calculated and defined by *IQVIA* as “the smallest common doses for a product form”, which “equates the number of mL of liquid preparations, such as 5 mL of codeine, to the standard solid dosage of one tablet”. *IQVIA’s* sample of data is based on audits, but their method of collection and coverage varies from country to country. Annual population statistics for each calendar year (2013 to 2018) were sourced from the World Bank [37].

### 2.2 Data analysis

We extracted details from the pack information and used descriptive statistics to determine the numbers and types of products sold across the 31 countries. Data on dosages were missing from the pack information for most countries, so we could not calculate oral morphine equivalents (OME), as this conversion would vary for the different products in each country. We combined the quarterly data to calculate the total dosage units sold over the study period and the totals for each year (e.g. from quarter two in 2013 to quarter one in 2014). We also calculated the mean number of dosage units sold over six years, adjusted for population. We created an annual rate of dosage units sold per 1000 of each year’s population for each country to examine trends over time. To adjust for the heterogeneity of *IQVIA*’s coverage by dividing the reported sales by the percentages in Table S1 of Supplement.

For public expenditure, *IQVIA* converted sales to pounds sterling (GBP, £) for each country on the date of data extraction (16 September 2019). Public expenditure refers to the money spent on OTC products by citizens directly from their pockets and was defined by *IQVIA* as the “pharmacy selling price or consumer purchase price or price to the public”. We calculated annual totals, mean public expenditure for each country, adjusted for population, and a rate of GBP per 1000 population to assess changes over time. We also adjusted for the heterogeneity of *IQVIA*’s coverage by dividing the reported expenditure by the percentages in Table S1 of Supplement.

### 2.3 Software and data sharing

We used Stata v16 and Python v3 in Jupyter Notebooks with pandas [38], seaborn [39], and matplotlib [40] libraries for analysis and figures. The information provided by *IQVIA* is considered commercial and requires a fee to access. Thus, we cannot openly share the data owing to contractual agreements with *IQVIA*. However, we have openly shared our statistical code at GitHub [41], preregistered [42] and published [43] our study protocol, and shared all our study materials via the Open Science Framework (OSF) [44].

## 3 Results

31.5 billion dosage units of codeine reportedly sold OTC across 31 countries over the six-year study period (April 2013 to March 2019). After adjusting for *IQVIA’s* data coverage, this equated to 42.8 billion dosage units. However, the distribution of reported sales was not uniform. Five countries represented 90% of OTC codeine sales reported by IQVIA; South Africa accounted for the greatest volume of sales data (34%), followed by France (20%), Japan (16.5%), the UK (14.5%), and Poland (5%).

South Africa sold the most OTC codeine products (mean of 36 dosage units/person; Figure 1 and Table 1), followed by Ireland (30 dosage units/person), France (20 dosage units/person), the UK (17.2 dosage units/person), and Latvia (16.8 dosage units/person).

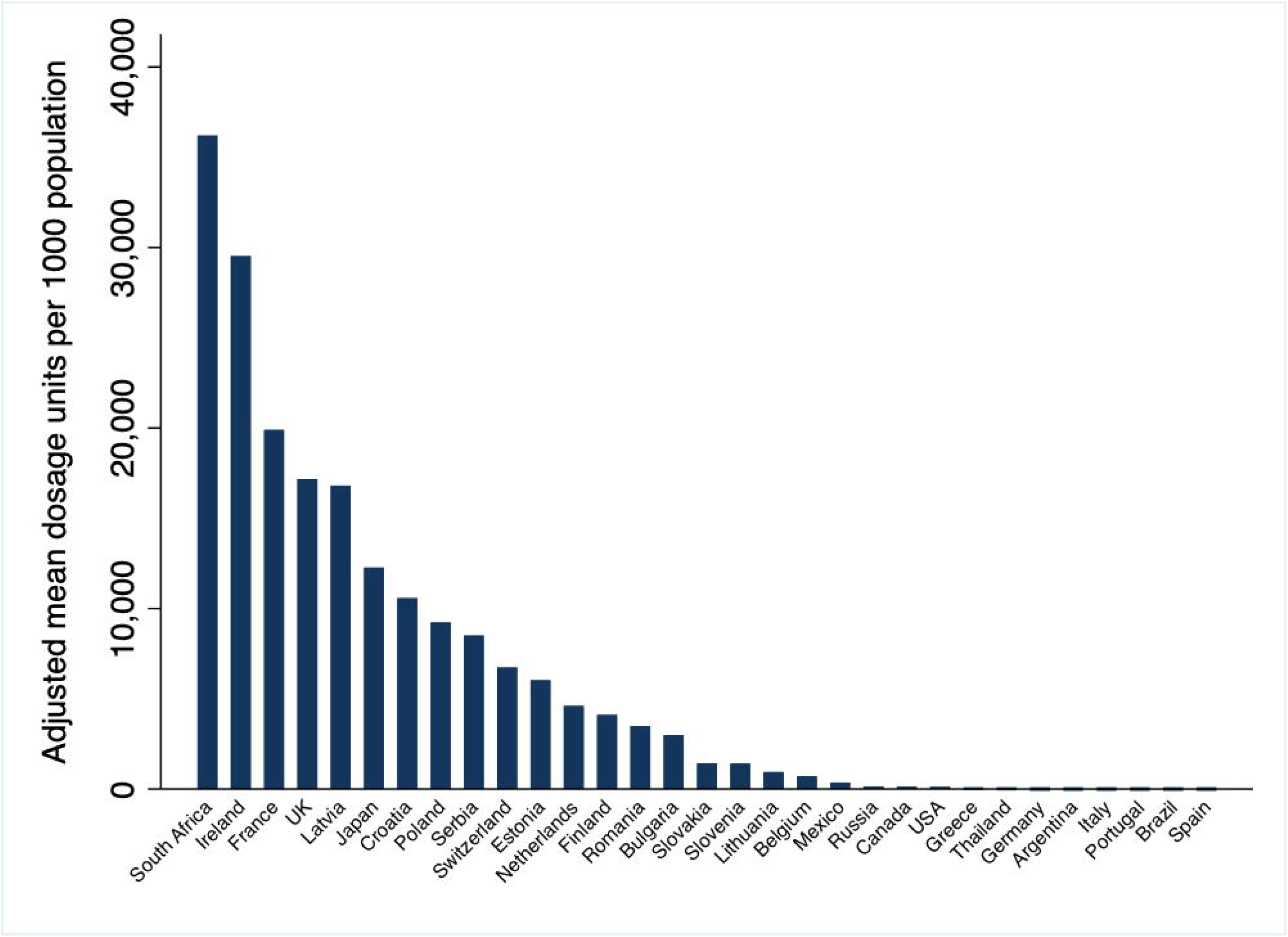

**Table 1:**
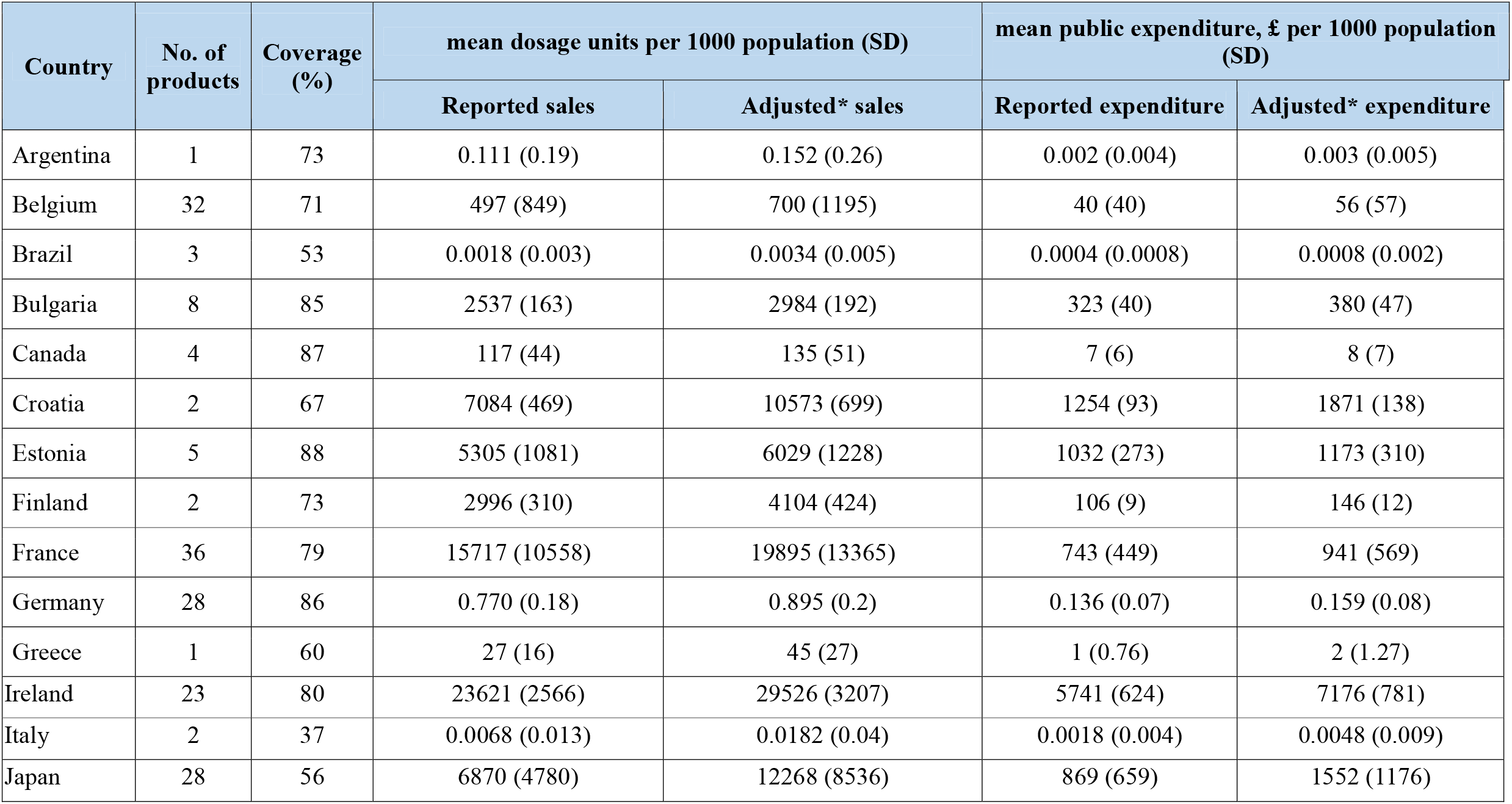
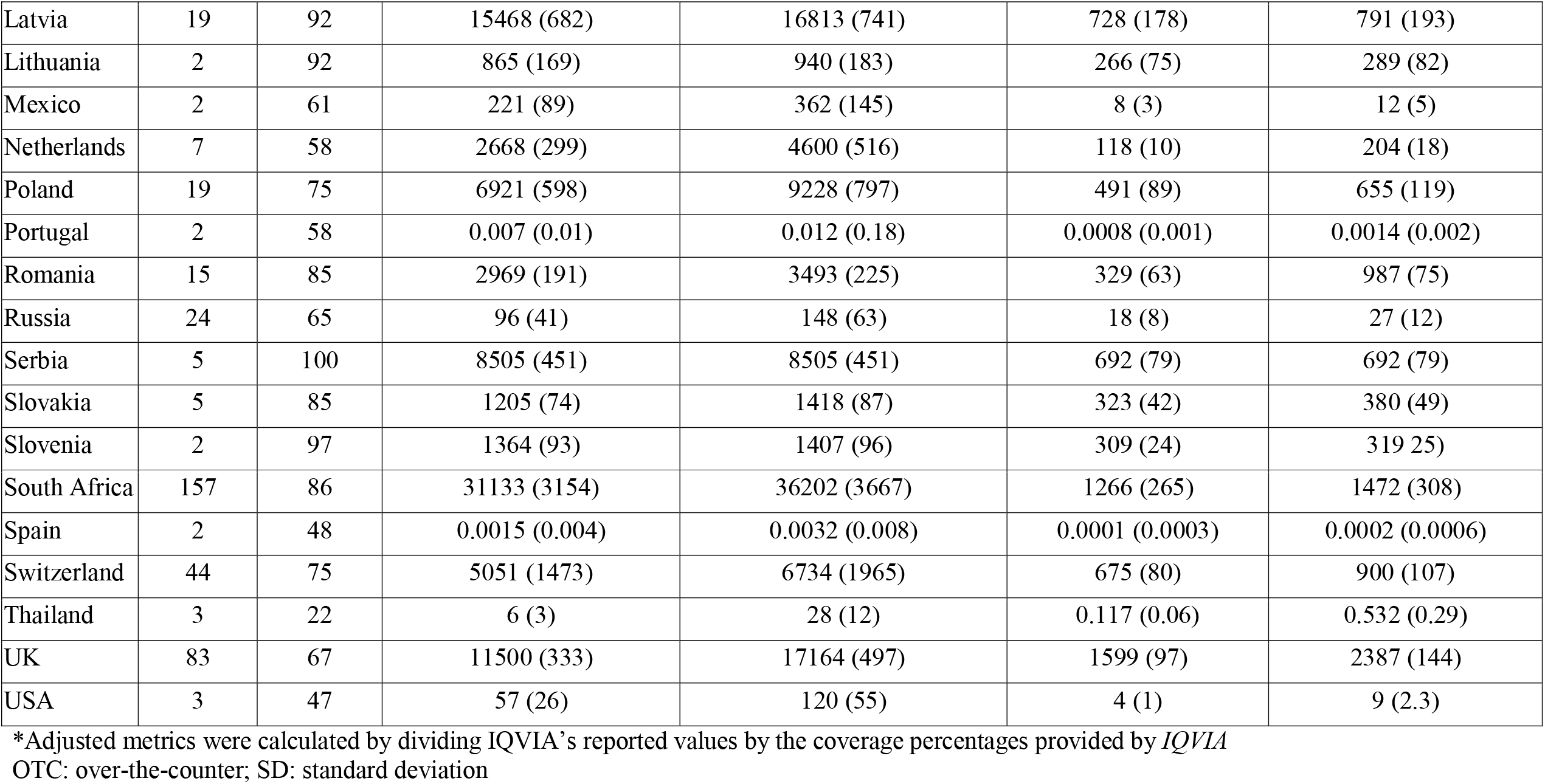
Mean sales of OTC codeine-containing products and public expenditure reported by *IQVIA* between April 2013 and March 2019 in 31 countries, and adjusted for *IQVIA’s* data coverage, in alphabetical order

| Country | No. of products | Coverage (%) | mean dosage units per 1000 population (SD) |  | mean public expenditure, £ per 1000 population (SD) |  |
| --- | --- | --- | --- | --- | --- | --- |
|  |  |  | Reported sales | Adjusted* sales | Reported expenditure | Adjusted* expenditure |
| Argentina | 1 | 73 | 0.111 (0.19) | 0.152 (0.26) | 0.002 (0.004) | 0.003 (0.005) |
| Belgium | 32 | 71 | 497 (849) | 700 (1195) | 40 (40) | 56 (57) |
| Brazil | 3 | 53 | 0.0018 (0.003) | 0.0034 (0.005) | 0.0004 (0.0008) | 0.0008 (0.002) |
| Bulgaria | 8 | 85 | 2537 (163) | 2984 (192) | 323 (40) | 380 (47) |
| Canada | 4 | 87 | 117 (44) | 135 (51) | 7 (6) | 8 (7) |
| Croatia | 2 | 67 | 7084 (469) | 10573 (699) | 1254 (93) | 1871 (138) |
| Estonia | 5 | 88 | 5305 (1081) | 6029 (1228) | 1032 (273) | 1173 (310) |
| Finland | 2 | 73 | 2996 (310) | 4104 (424) | 106 (9) | 146 (12) |
| France | 36 | 79 | 15717 (10558) | 19895 (13365) | 743 (449) | 941 (569) |
| Germany | 28 | 86 | 0.770 (0.18) | 0.895 (0.2) | 0.136 (0.07) | 0.159 (0.08) |
| Greece | 1 | 60 | 27 (16) | 45 (27) | 1 (0.76) | 2 (1.27) |
| Ireland | 23 | 80 | 23621 (2566) | 29526 (3207) | 5741 (624) | 7176 (781) |
| Italy | 2 | 37 | 0.0068 (0.013) | 0.0182 (0.04) | 0.0018 (0.004) | 0.0048 (0.009) |
| Japan | 28 | 56 | 6870 (4780) | 12268 (8536) | 869 (659) | 1552 (1176) |
| Latvia | 19 | 92 | 15468 (682) | 16813 (741) | 728 (178) | 791 (193) |
| Lithuania | 2 | 92 | 865 (169) | 940 (183) | 266 (75) | 289 (82) |
| Mexico | 2 | 61 | 221 (89) | 362 (145) | 8 (3) | 12 (5) |
| Netherlands | 7 | 58 | 2668 (299) | 4600 (516) | 118 (10) | 204 (18) |
| Poland | 19 | 75 | 6921 (598) | 9228 (797) | 491 (89) | 655 (119) |
| Portugal | 2 | 58 | 0.007 (0.01) | 0.012 (0.18) | 0.0008 (0.001) | 0.0014 (0.002) |
| Romania | 15 | 85 | 2969 (191) | 3493 (225) | 329 (63) | 987 (75) |
| Russia | 24 | 65 | 96 (41) | 148 (63) | 18 (8) | 27 (12) |
| Serbia | 5 | 100 | 8505 (451) | 8505 (451) | 692 (79) | 692 (79) |
| Slovakia | 5 | 85 | 1205 (74) | 1418 (87) | 323 (42) | 380 (49) |
| Slovenia | 2 | 97 | 1364 (93) | 1407 (96) | 309 (24) | 319 (25) |
| South Africa | 157 | 86 | 31133 (3154) | 36202 (3667) | 1266 (265) | 1472 (308) |
| Spain | 2 | 48 | 0.0015 (0.004) | 0.0032 (0.008) | 0.0001 (0.0003) | 0.0002 (0.0006) |
| Switzerland | 44 | 75 | 5051 (1473) | 6734 (1965) | 675 (80) | 900 (107) |
| Thailand | 3 | 22 | 6 (3) | 28 (12) | 0.117 (0.06) | 0.532 (0.29) |
| UK | 83 | 67 | 11500 (333) | 17164 (497) | 1599 (97) | 2387 (144) |
| USA | 3 | 47 | 57 (26) | 120 (55) | 4 (1) | 9 (2.3) |

\*Adjusted metrics were calculated by dividing IQVIA's reported values by the coverage percentages provided by *IQVIA* OTC: over-the-counter; SD: standard deviation

Over the six-year study period, total reported sales increased by 2.8%, from 3025 dosage units/1000 population in 2013–14 to 3111 dosage units/1000 in 2018–19. After adjusting for *IQVIA’s* data coverage, total sales increased by 11.4%, from 3911 dosage units/1000 population in 2013–14 to 4358 dosage units/1000 in 2018-19. Nationally, most (52%, n=16) countries had decreased OTC codeine sales, but this varied widely (Figure 2, and Figure S1 and S2 in Supplement 1).

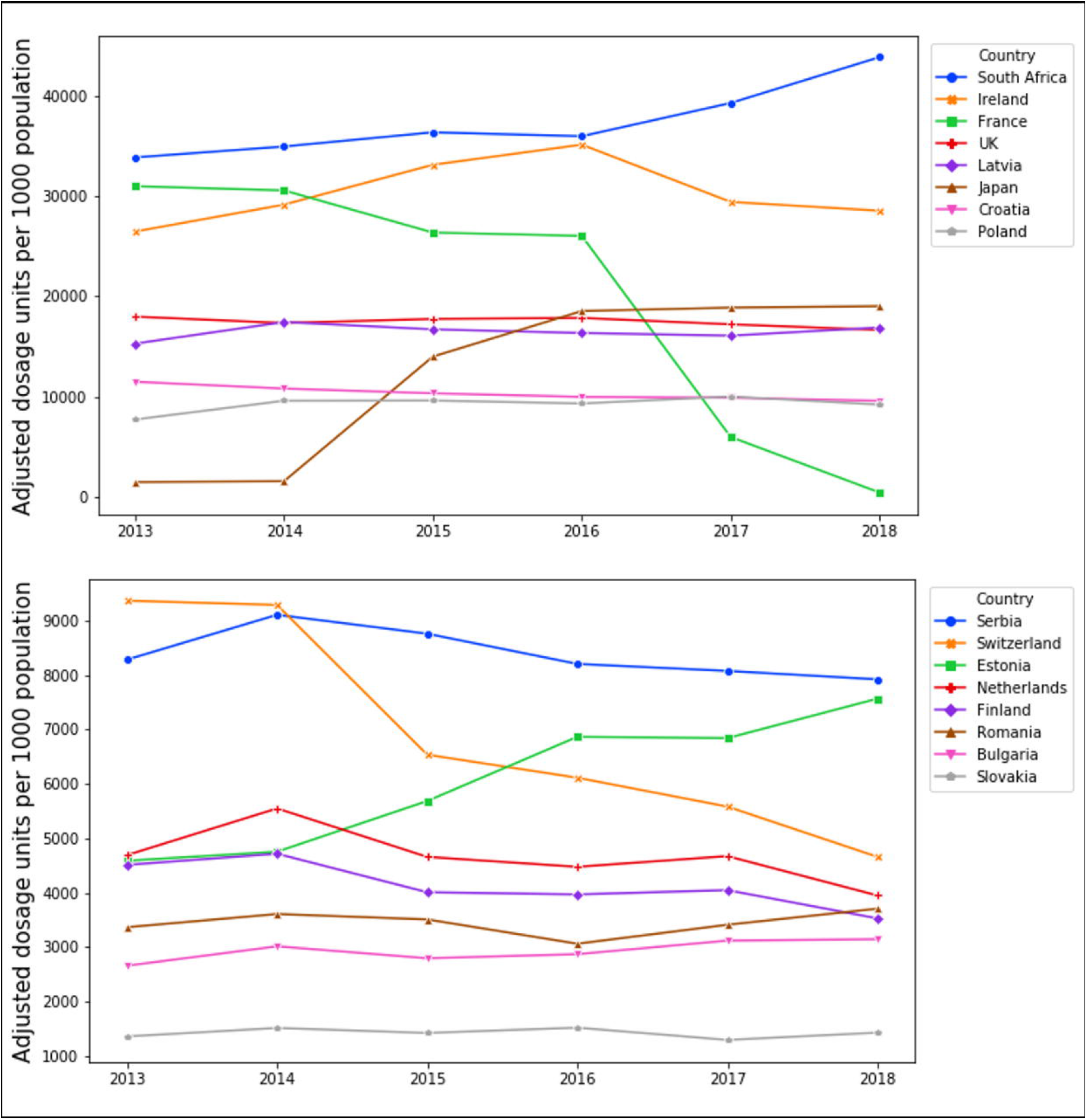

The public spent £2.55 billion on OTC codeine-containing products in 31 countries over six years; after adjusting *IQVIA’s* percentage coverage, this equated to £3.68 billion. Reported public expenditure increased by 54%, from £196/1000 population in 2013–14 to £301/1000 population in 2018–19. Adjusting for *IQVIA’s* percentage coverage, public expenditure increased by 72%, from £263/1000 population in 2013–14 to £451/1000 population in 2018–19.

Ireland had the largest mean public expenditure of £7.18 per person, followed by the UK (mean of £2.39/person), Croatia (£1.87/person), Japan (£1.55/person), and South Africa (£1476/person) (Figure 3).

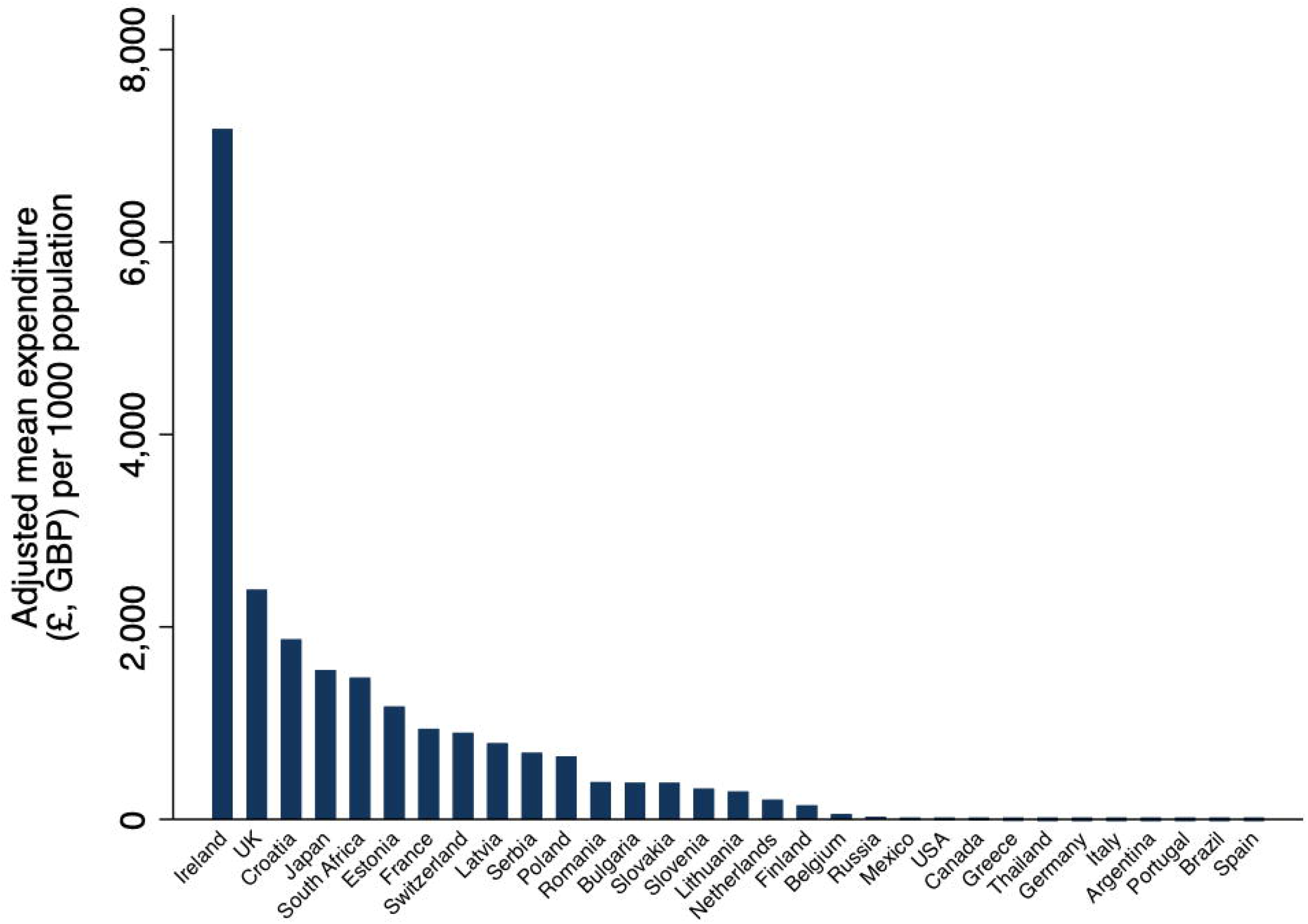

Most countries (58%, 18/31) had increased public expenditure over time. There were simultaneous increases (45%, 14/31) and decreases (39%, 12/31) in both sales and expenditure in most countries, while other countries (16%, 5/31) had a discordance in the direction of their sales and expenditure (Figure 4).

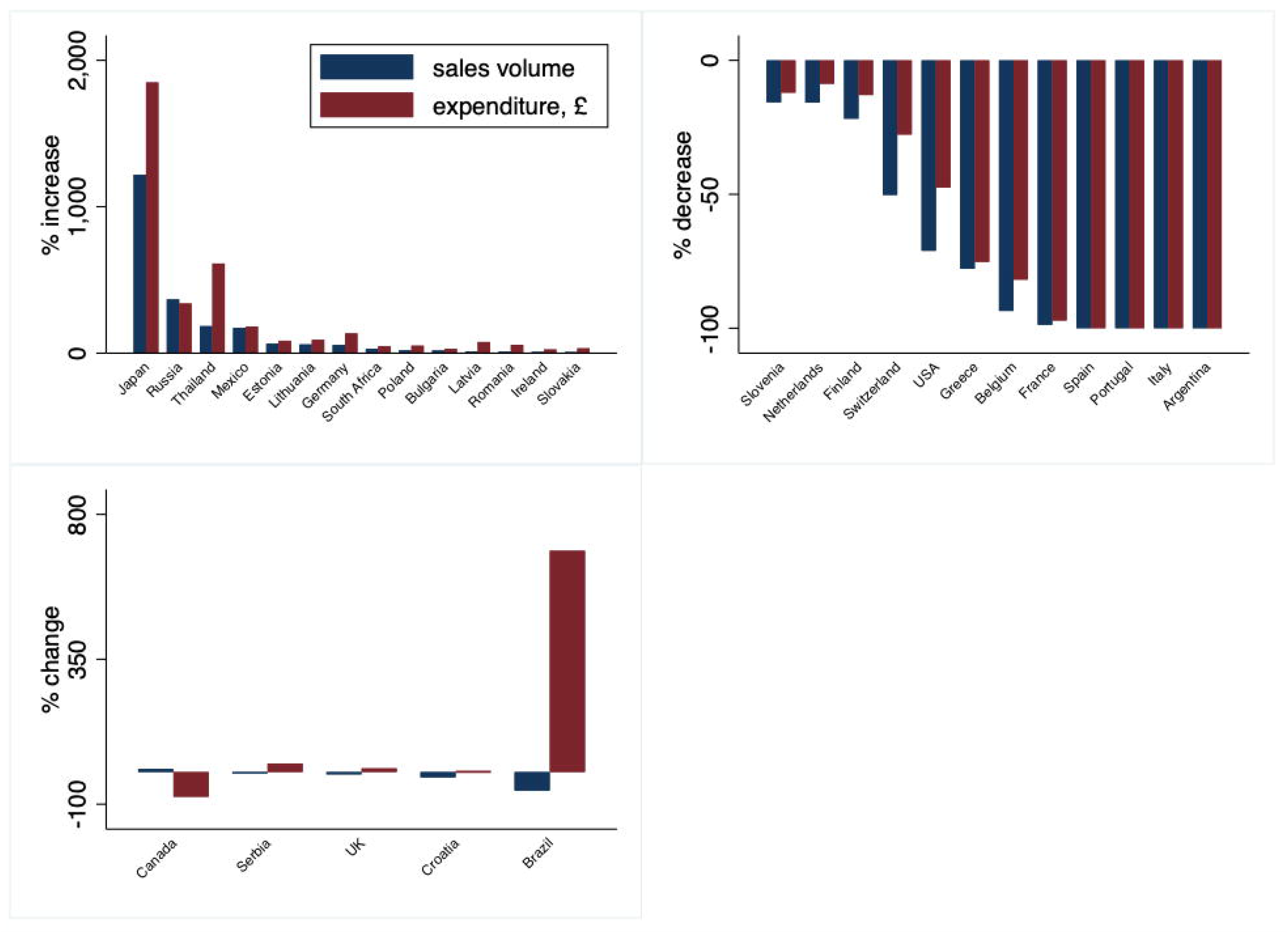

There were 569 products and 12 formulations sold across 31 countries (Figure S3 in Supplement 1). Tablets (40%) were the most common formulations sold, followed by syrups (22%), soluble tablets (9%), and coated tablets (8%). Seven countries, including Argentina, Finland, Greece, Italy, Mexico, The Netherlands, and The USA, reportedly sold no codeine products in any tablet formulations OTC; instead, they sold codeine in liquid, syrup, lozenge, drop or powder formulations (Table S2 in Supplement 1). Products contained a median of three substances per combination (IQR: 2–4, range: 1– 16). Limited details were available in the pack information: the dosages of codeine were available in 17% of products (98 of 569) in 15 countries.

## 4 Discussion

Many people have purchased non-prescribed codeine in several parts of the world. Overall, total sales and public expenditure of OTC codeine products increased over time. However, sales and expenditure were not equally distributed across the 31 countries.

South Africa consistently sold the greatest volume of OTC codeine each year. This demand for OTC codeine could be driven by the incidences of painful conditions in South Africa and the growing concern of opioid misuse and dependence [45,46]. In the 2016 South African Demographic and Household Survey [45], the prevalence of chronic pain in the adult population was 18.3% (95% confidence interval: 17.0-19.7%). A study of admission for substance abuse treatments in South Africa found that 2.5% (n=435) of all admissions in 2014 were for codeine misuse or dependence [47]. In a 2014 survey of prescribers in South Africa, concerns were expressed about the availability of codeine OTC and the lack of data sources to examine its use [46]. The availability of non-prescribed codeine in South Africa may also have ramifications for neighbouring countries that restrict access. For example, reports in Zimbabwe suggest that codeine-containing cough syrup was being illegally smuggled in from South Africa and sold on the streets after being outlawed in 2015 [48]. However, there are limited data in many countries on the prevalence of such activities and the extent of codeine use and misuse.

The public in Ireland spent the most on OTC codeine products and had the second-largest sales volume. Concerns regarding public access to codeine and its misuse are well reported in Ireland [49,50]. However, a study examining hospital presentations involving intentional drug overdoses between 2007 and 2013 in Ireland reported a 20% decrease in codeine-related overdoses following new guidance for pharmacists in 2010 [51]. Despite this guidance, the sales of codeine in Ireland remained high in our study.

According to *IQVIA*’s data, Japan had the largest increase in sales, which may have been partly driven by Japan’s reclassification process for OTC medicines in 2015 [52]. In contrast, sales fell considerably in France, which can be attributed to the decree signed by the French government in July 2017 that made codeine prescription-only with immediate effect [27]. In South Africa, Canada, Switzerland, Ireland, and the UK, governments have proposed or are considering plans to do likewise [53–56], but in the meantime, many people in these countries may be self-medicating and unknowingly developing codeine dependency or addiction. Studies assessing the effect of rescheduling codeine to prescription-only in Australia showed a reduction in all codeine-related poisonings and no change in calls to poisons centres or sales of high-strength (>15 mg) prescribed codeine after reclassification [31,57]. The success of Australia’s rescheduling suggests that governments worldwide should make codeine prescription-only. However, since many low- and middle-income countries experience barriers to accessing opioids [58–60], the WHO recommends that codeine should not be rescheduled and for codeine to be included in essential medicines lists [61]. If a consensus on the status of OTC codeine products cannot be reached, data should be collected globally to monitor its use and harms.

Changes to regulations of OTC codeine and differences in trade exemptions, agreements, and disclosures of commercial interests at the country level may explain some of the variations in sales and expenditure [25]. For example, the public in Ireland, the UK, Croatia, and Japan spent more on OTC codeine products than South Africa despite their large sales volume. In countries such as Germany and the USA, which had high rates of prescribed opioids [60], mean sales of OTC codeine products were low. However, our figures included various formulations of codeine available OTC and depended on the coverage of data from *IQVIA* during the study period. Thus, it is difficult to determine whether the variation in sales represents real differences between countries.

### Strengths and limitations

We preregistered our study protocol and shared our statistical code and study materials where possible [41,42,44]. We used dosage units to combine liquid and solid forms, accounted for population increases over the study period and adjusted for *IQVIA’s* coverage of data. The figures represent population-level sales and expenditure of OTC codeine in 31 countries, providing the best available proxy for actual use. However, many other countries sold codeine OTC during our study period, including Australia [32], which was not provided by *IQVIA* when we requested the data. Due to such data’s commercial nature, *IQVIA* also withheld methodological information about the data, such as their formulation for calculating dosage units and conversion to GBP for other currencies. *IQVIA*’s coverage and the completeness of data may have also affected trends; the percentages on data coverage was provided at single time points, which may not accurately represent changes over time. Sales represented products for adults, although we calculated rates using population statistics for all age groups, including children. Codeine-containing products may also be purchased in large quantities from online pharmacies or the black market [62], not captured in these data.

### Implications

Since access to data on OTC codeine sales has previously been difficult to assess, our study provides one of the first estimates of the amount of codeine sold OTC in the 31 included countries. This information could inform future reviews of codeine by the WHO’s Expert Committee on Drug Dependence [25] and international policies such as the rescheduling of codeine in the 1961 Single Convention on Narcotics Drugs.

However, better access to OTC sales data is still required. Amendments to medicines legislation in the UK shows how such data could be collected. The Misuse of Drugs Regulation 2001, the Medicines for Human Use (Administration and Sale or Supply) (Miscellaneous Amendments) Order 2007, and the Medicines (Sale or Supply) (Miscellaneous Provisions Amendment Regulations 2007) were updated to require pharmacies to submit counts of private prescriptions for Schedule 2 and Schedule 3 controlled drugs to the National Health Service (NHS) Prescription Services for analysis, audit, and monitoring [63,64]. A similar system could be enforced through a public health organisation such as the WHO or the International Narcotics Control Board (INCB), which already collects governments’ annual drug statistics on codeine [65]. Such data could then be used by governments and researchers to monitor sales of OTC codeine and measure the impact of regulatory changes.

### Conclusions

Codeine is one of the most widely accessible and used opioids worldwide. However, monitoring its use and preventing its misuse as an OTC product is a public health challenge. Healthcare professionals should ask their patients about their use of OTC products. Public health measures are needed to identify and prevent codeine misuse and increase awareness and education of the harms of codeine, particularly in young adults. Governments should review policies to improve the collection of sales data, and the safety of products sold OTC containing codeine.

## Data Availability

Study materials, protocol, and statistical code are openly available on our OSF repository [64] (https://osf.io/yt6bf/) and at GitHub [61] (https://github.com/georgiarichards/otc_codeine). We cannot openly share the data owing to contractual agreements with IQVIA, but the data can be accessed directly from IQVIA, which will require a fee.

https://github.com/georgiarichards/otc_codeine

## Acknowledgements

We thank the Primary Care Research Trust of Birmingham and Midlands Research Practices Consortium for providing the funding to purchase the sales data from IQVIA.

## Supplementary material

Supplement 1: Supplementary tables and figures

Supplement 2: STROBE reporting checklist

## Declarations

### Funding

This research was supported by the Primary Care Research Trust of Birmingham and Midlands Research Practices Consortium who provided the funding to purchase the sales data from IQVIA.

### Competing interests

GCR was financially supported by the National Institute for Health Research (NIHR) School for Primary Care Research (SPCR), the Naji Foundation, and the Rotary Foundation to study for a Doctor of Philosophy (2017-2020), but no longer has any financial COIs. GCR is an Associate Editor of BMJ Evidence Based Medicine. JKA has published articles and edited textbooks on adverse drug reactions and interactions and has often given medicolegal advice, including appearances as an expert witness in coroners’ courts, often dealing with the adverse effects of opioids. BM works for NHS England as a pharmacist adviser. BG has received research funding from the Laura and John Arnold Foundation, the NIHR, the NIHR SPCR, the NIHR Oxford Biomedical Research Centre, the Mohn-Westlake Foundation, NIHR Applied Research Collaboration Oxford and Thames Valley, the Wellcome Trust, the Good Thinking Foundation, Health Data Research UK (HDR UK), the Health Foundation, and the World Health Organisation (WHO); he also receives personal income from speaking and writing for lay audiences on the misuse of science. FDRH acknowledges part support from the NIHR SPCR, the NIHR Applied Research Collaboration (ARC) Oxford Thames Valley, and the NIHR Oxford OUH BRC. CH is an NIHR Senior Investigator and has received expenses and fees for his media work, expenses from the WHO, FDA, and holds grant funding from the NIHR SPCR and the NIHR SPCR Evidence Synthesis Working Group [Project 380], the NIHR BRC Oxford and the WHO. On occasion, CH receives expenses for teaching EBM and is also paid for his GP work in NHS out of hours (contract with Oxford Health NHS Foundation Trust). CH is the Director of the CEBM. The views expressed are those of the authors and not necessarily those of the NHS, the NIHR or the Department of Health and Social Care.

### Availability of data and material

Study materials are available on an open repository [44] (https://osf.io/yt6bf/). We cannot openly share the data owing to contractual agreements with *IQVIA*, but the data can be accessed directly from *IQVIA*, which will require a fee.

### Code availability

Our statistical code is openly available at GitHub [41]

(https://github.com/georgiarichards/otc_codeine).

### Authors’ contributions

GCR devised the research question, designed the methods, wrote the protocol, conducted a literature search, sourced the data, cleaned, managed, and analysed the data, created the figures, and wrote the first draft of the manuscript. JKA and CH reviewed the protocol and preliminary findings and provided supervisory support. FDRH reviewed the protocol and facilitated the grant application. BM reviewed preliminary findings and contributed to the interpretation of data. BG provided supervisory support. All authors read and approved the final version.

### Ethics approval

Not applicable

### Consent to participate

Not applicable

### Consent to publication

All authors read and approved the final manuscript and consent to submit the manuscript for publication.

## Notes

### Clinical Trial

https://osf.io/ay4mc

### Clinical Protocols

https://doi.org/10.17605/OSF.IO/AY4MC

### Funding Statement

The Primary Care Research Trust of Birmingham and Midlands Research Practices Consortium provided the funds to purchase the sales data from IQVIA. The sponsor was not involved in the study design, collection, analysis or interpretation of data, writing of the manuscript, or decisions on submitting the manuscript for publication. The researchers had full access to all the data used in this study, which was independent from the funders. The authors take responsibility for the integrity of the data and the accuracy of the data analysis.

### Summary of Updates

Revised manuscript from peer review comments

