## Supplementary material for "Sales of over-the-counter products containing codeine in 31 countries, 2013-2019: a retrospective observational study": Table S1 in Supplement 1

**Journal**: Drug Safety

**Authors:** *Georgia C Richards* (0000-0003-0244-5620)^1,2^; Jeffrey K Aronson (0000-0003-1139-655X)^2^; Brian MacKenna (0000-0002-3786-9063)^3^; Ben Goldacre (0000-0002-5127-4728)^3^; FD Richard Hobbs (0000-0001-7976-7172)^4^; Carl Heneghan (0000-0002-1009-1992)^1,2^*

^1^Global Centre on Healthcare and Urbanisation, Kellogg College, University of Oxford, 60-62 Banbury Road, Oxford, OX2 6PN, UK.

^2^Centre for Evidence-Based Medicine, Nuffield Department of Primary Care Health Sciences, University of Oxford, Radcliffe Observatory Quarter, Woodstock Road, Oxford, OX2 6GG, UK.

^3^EBM Datalab, Nuffield Department of Primary Care Health Sciences, University of Oxford, Radcliffe Observatory Quarter, Woodstock Road, Oxford, OX2 6GG, UK.

^4^Nuffield Department of Primary Care Health Sciences, University of Oxford, Radcliffe Observatory Quarter, Woodstock Road, Oxford, OX2 6GG, UK.

**Table S1**: Coverage of *IQVIA*’s data by country in descending order

| **country** | **% of the total market** |
| --- | --- |
| Serbia | 100 |
| Slovenia | 97 |
| Latvia | 92 |
| Lithuania | 92 |
| Estonia | 88 |
| Canada | 87 |
| Germany | 86 |
| South Africa | 86 |
| Bulgaria | 85 |
| Romania | 85 |
| Slovakia | 85 |
| Ireland | 80 |
| France | 79 |
| Poland | 75 |
| Switzerland | 75 |
| Argentina | 73 |
| Finland | 73 |
| Belgium | 71 |
| Croatia | 67 |
| UK | 67 |
| Russia | 65 |
| Mexico | 61 |
| Greece | 60 |
| Netherlands | 58 |
| Portugal | 58 |
| Japan | 56 |
| Brazil | 53 |
| Spain | 48 |
| USA | 47 |
| Italy | 37 |
| Thailand | 22 |

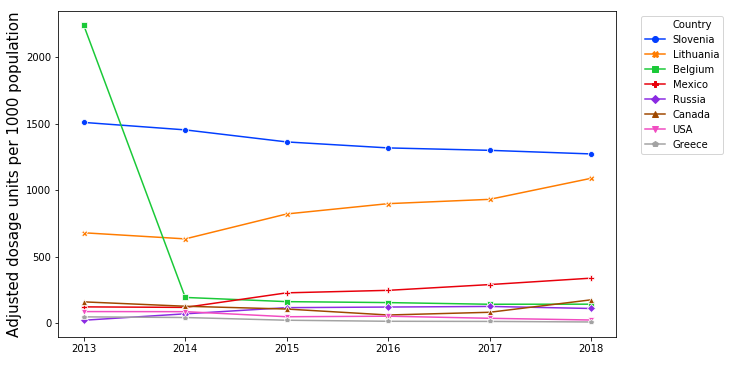

**Figure S1:** Over-the-counter products containing codeine sold per 1000 population starting in April 2013-March 2014, and ending in April 2018-March 2019, for countries in the third quartile of sales, based on their mean dosage units sold per person, adjusted for data coverage.

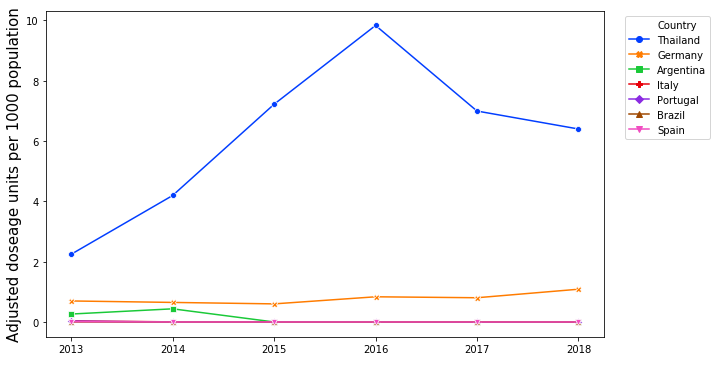

**Figure S2:** Over-the-counter products containing codeine sold per 1000 population starting in April 2013-March 2014, and ending in April 2018-March 2019, for countries in the last quartile of sales, based on their mean dosage units sold per person, adjusted for data coverage.

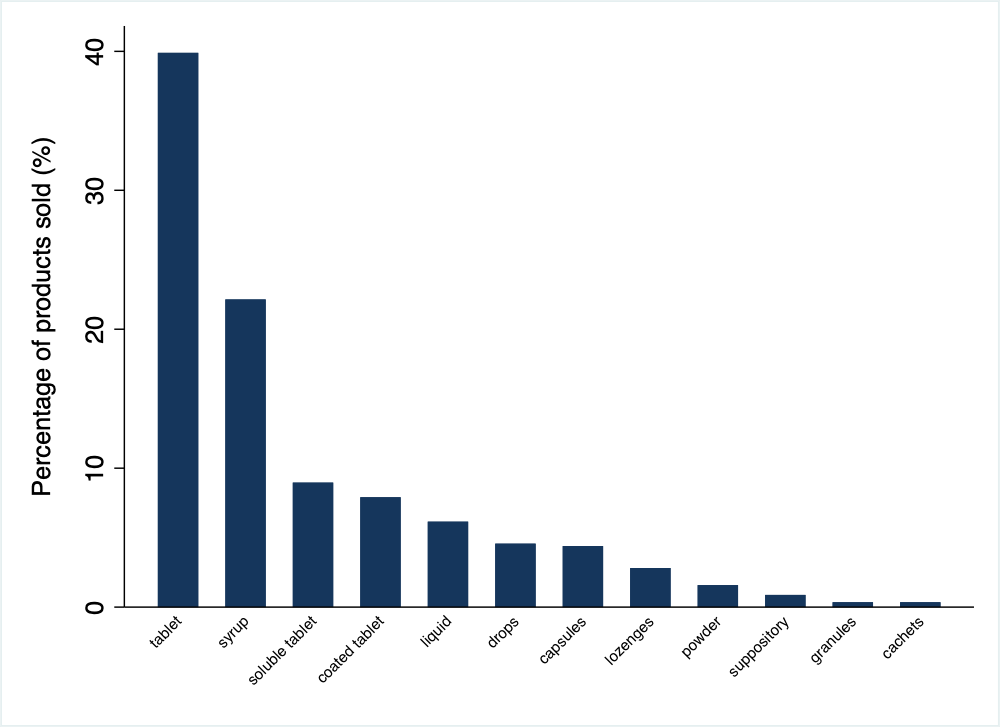

**Figure S3:** Percentage of OTC codeine-containing products sold by formulation for 31 countries over six years (April 2013 to March 2019).

**Table S2:** Frequency of the types of codeine formulations sold over-the-country by country in alphabetical order

| **Country** | **Tablet** | **Syrup** | **Soluble tablet** | **Coated tablets** | **Liquid** | **Drops** | **Capsules** | **Lozenges** | **Powder** | **Suppository** | **Granules** | **Cachets** |
| --- | --- | --- | --- | --- | --- | --- | --- | --- | --- | --- | --- | --- |
| Argentina |  | 1 |  |  |  |  |  |  |  |  |  |  |
| Belgium | 5 | 19 |  |  |  |  | 2 | 1 | 1 | 2 |  | 2 |
| Brazil | 2 |  |  |  | 1 |  |  |  |  |  |  |  |
| Bulgaria | 5 |  | 2 |  |  |  | 1 |  |  |  |  |  |
| Canada | 3 |  |  |  |  |  |  | 1 |  |  |  |  |
| Croatia | 2 |  |  |  |  |  |  |  |  |  |  |  |
| Estonia | 3 |  | 2 |  |  |  |  |  |  |  |  |  |
| Finland |  |  |  |  | 2 |  |  |  |  |  |  |  |
| France | 14 | 12 |  | 3 | 3 |  | 2 | 1 |  | 1 |  |  |
| Germany | 8 |  |  |  |  | 20 |  |  |  |  |  |  |
| Greece |  |  |  |  |  |  |  | 1 |  |  |  |  |
| Ireland | 7 |  | 5 | 6 | 3 |  | 2 |  |  |  |  |  |
| Italy |  |  |  |  |  | 2 |  |  |  |  |  |  |
| Japan | 9 |  |  | 3 | 7 |  | 2 |  | 5 |  | 2 |  |
| Latvia | 13 | 3 | 2 | 1 |  |  |  |  |  |  |  |  |
| Lithuania | 1 |  | 1 |  |  |  |  |  |  |  |  |  |
| Mexico |  | 1 |  |  |  |  |  |  | 1 |  |  |  |
| Netherlands |  | 6 |  |  | 1 |  |  |  |  |  |  |  |
| Poland | 8 | 5 | 3 | 2 |  |  | 1 |  |  |  |  |  |
| Portugal | 1 |  |  |  |  |  |  | 1 |  |  |  |  |
| Romania | 9 |  | 2 | 4 |  |  |  |  |  |  |  |  |
| Russia | 20 | 1 | 2 |  |  |  | 1 |  |  |  |  |  |
| Serbia | 4 | 1 |  |  |  |  |  |  |  |  |  |  |
| Slovakia | 1 |  | 2 |  |  | 1 | 1 |  |  |  |  |  |
| Slovenia | 2 |  |  |  |  |  |  |  |  |  |  |  |
| South Africa | 84 | 53 | 4 | 4 | 6 |  | 6 |  |  |  |  |  |
| Spain | 1 |  |  |  | 1 |  |  |  |  |  |  |  |
| Switzerland | 1 | 22 |  | 2 |  | 3 | 2 | 11 | 1 | 2 |  |  |
| Thailand | 3 |  |  |  |  |  |  |  |  |  |  |  |
| UK | 21 |  | 26 | 20 | 11 |  | 5 |  |  |  |  |  |
| USA |  | 2 |  |  |  |  |  |  | 1 |  |  |  |
